# *De novo* protein-coding gene variants in developmental stuttering

**DOI:** 10.1101/2024.11.25.24317778

**Authors:** Else Eising, Ivana Dzinovic, Arianna Vino, Lottie Stipdonk, Martin Pavlov, Juliane Winkelmann, Martin Sommer, Marie-Christine J.P. Franken, Konrad Oexle, Simon E. Fisher

## Abstract

Stuttering is a common neurodevelopmental condition characterized by disfluencies in speech, such as blocks, prolongations, and repetitions. While most children who stutter do so only transiently, there are some for whom stuttering persists into adulthood. Rare-variant screens in families including multiple relatives with persistent stuttering have so far identified six genes carrying putative pathogenic variants hypothesized to act in a monogenic fashion. Here, we applied a complementary study design, searching instead for *de novo* variants in exomes of 85 independent parent-child trios, each with a child with transient or persistent stuttering. Exome sequencing analysis yielded a pathogenic variant in *SPTBN1* as well as likely pathogenic variants in *PRPF8*, *TRIO*, and *ZBTB7A* - four genes previously implicated in neurodevelopmental disorders with or without speech problems. Our results also highlighted two further genes of interest for stuttering: *FLT3* and *IREB2*. We used extensive bioinformatic approaches to investigate overlaps in brain-related processes among the twelve genes associated with monogenic forms of stuttering. Analyses of gene-expression datasets of the developing and adult human brain, and data from a genome-wide association study of human brain structural connectivity, did not find links of monogenic stuttering to specific brain processes. Overall, our results provide the first direct genetic link between stuttering and other neurodevelopmental disorders, including speech delay and aphasia. In addition, we systematically demonstrate a dissimilarity in biological pathways associated with the genes thus far implicated in monogenic forms of stuttering, indicating heterogeneity in the etiological basis of this condition.

## Introduction

Developmental stuttering is characterized by disfluencies in speech, such as blocks, prolongations and repetitions. It generally starts early in childhood, between 2 and 5 years of age, affecting approximately 8% of children^1^. While the majority of children recover naturally or with speech therapy within a few years, stuttering persists in a subset of individuals, leading to persistent stuttering in approximately 0.8% of the population^2^, three to four times as often in men than in women^1,3^. In adults, moments of speech dysfluency are usually accompanied by various cognitive, behavioural and emotional reactions, which often become a central component of stuttering^4^. Through these internal reactions and negative feedback from the environment, stuttering can have a major impact on a person’s physical, psychological, and social quality of life^5^.

It is well established that genetic factors play a role in the development of stuttering. Large twin studies on stuttering found a heritability of 40% to 80%^6–8^. Three genome-wide association studies identified the first genome-wide significant loci associated with stuttering^9–11^, indicating that this is a genetically complex multifactorial trait for at least some of the affected population. In addition, the inheritance patterns observed in some large families suggest that stuttering may sometimes occur as a Mendelian (monogenic) trait, involving a single rare gene variant with a large effect size. Rare variant screens using linkage analysis followed by Sanger sequencing, and more recently using next generation sequencing, identified six genes to be associated with persistent stuttering in large families: *GNPTAB*^12^*, AP4E1*^13^*, IFNAR1*^14^*, ARMC3*^15^*, ZBTB20*^16^ and *PPID*^17^. Hypothesis-driven genetic screens in unrelated people who stutter and controls also suggested an increased burden of rare variants in *GNPTG* and *NAGPA*, two genes that function in the same enzymatic pathway as *GNPTAB*^12,18^. The aetiology of stuttering may therefore involve both complex polygenic and monogenic causal factors, but the relative contributions of these different types of genetic influence is not yet known.

The genes implicated in monogenic forms of stuttering have so far not pointed to a shared biological mechanism, but instead are involved in a wide variety of cellular functions. *GNPTAB, GNPTG* and *NAGPA* encode enzymes that synthesize mannose 6-phosphate recognition markers onto lysosomal enzymes^19^. *AP4E1* encodes a subunit of an adaptor protein involved in intracellular trafficking of vesicles of the Golgi, trans-Golgi network and endosomes, and is hypothesized to control autophagy^20^. *IFNAR1* encodes a subunit of the interferon receptor IFNR that can be activated by type I interferons during pathogen infections and autoimmune reactions^21^. *ARMC3* expresses a protein containing armadillo repeats, which produces a distinct structure that facilitates protein-protein interactions^22^. The exact function of ARMC3 is unknown, but related proteins are often involved signal transduction and cytoskeleton regulation. *ZBTB20* encodes a transcription factor with essential roles in multiple organ systems^23^. And lastly, *PPID* encodes an import component of the mitochondrial permeability transition pore, a protein complex whose expression can lead to mitochondrial swelling and cell death through apoptosis^24^. Only a few lines of indirect evidence have pointed towards potential overlapping processes, besides the shared enzymatic pathway of *GNTPAB*, *GNPTG* and *NAGPA*. First, NAGPA and AP4E1 have been shown to interact in a yeast-two-hybrid system^13^. Second, variants in *GNPTAB* and *PPID* have been reported to affect white matter features in transgenic knock-in mouse models, as immunohistochemical staining of the Gfap astrocyte marker was decreased in the corpus callosum of *Gnptab* mouse model^25^, and the microstructure of the left corticospinal tract in the *Ppid* mouse model differed in a brain imaging analysis^17^. Other links between the genes implicated in monogenic stuttering are yet to be identified.

Overlap in disease mechanisms is also not evident from examining the other monogenic disorders that are caused by genes thus far implicated in stuttering. Homozygous loss of function of *GNPTAB* and *GNPTG* are a cause of mucolipidosis, a severe lysosomal storage disorder^26,27^. Homozygous mutations in *AP4E1* are a cause of hereditary spastic paraplegia, a neurodevelopmental disorder characterized by developmental delay, moderate to severe intellectual disability and neonatal hypotonia that progresses to spasticity^28^. Homozygous loss-of-function mutations in *IFNAR1* cause an immunologic disorder characterized by increased susceptibility to viral infections^28^. Lastly, heterozygous missense variants in *ZBTB20* cause Primrose syndrome, a neurodevelopmental disorder characterized by intellectual disability, macrocephaly, unusual facial features and progressive features such as hearing loss and muscle wasting^29^. Of note, the types of mutations associated with stuttering are different from the types of mutations associated with these other Mendelian disorders: mainly heterozygous missense variants have been associated with stuttering in *GNPTAB* and *GNPTG*^12,18^, *AP4E1*^13^ and *IFNAR1*^14^, while a homozygous missense variant was associated with stuttering in *ZBTB20*^16^. These other Mendelian disorders therefore do not yet help elucidate important biological processes involved in stuttering. In contrast, monogenic forms of childhood apraxia of speech^30–32^ and speech delay^33^ are caused by genes often implicated in neurodevelopmental disorders characterized by intellectual disability, autism and epilepsy through the same types of variants, that often have functions involved in gene expression regulation, and that show co-expression during early brain development. Identifying additional genes causal for monogenic forms of stuttering is hence important for increasing understanding of the underlying biological mechanisms.

The present study aimed to apply a novel strategy to identify genes involved in monogenic forms of stuttering, moving beyond the multiplex family approaches of prior work. We applied whole exome sequencing to 85 parent-offspring trios, each with a child who stutters or stuttered in the past, and two parents who never stuttered, and searched for *de novo* variants that were present in the DNA of the child but not in the DNA of both parents. This trio design has been highly successfully applied to other neurodevelopmental disorders^34^ as well as to childhood apraxia of speech^31,32,35^, but genetic research on stuttering has yet to make use of this. Next, we applied several *in silico* analyses to investigate whether genes associated with monogenic stuttering show overlap in brain-relevant biological functions involving brain development and white matter structure. Our work identified four new genes with (likely) pathogenic *de novo* variants and highlighted another two genes of interest for stuttering. In contrast with other neurodevelopmental disorders including childhood apraxia of speech, genes implicated in monogenic forms of stuttering show highly diverse expression patterns in the developing brain and the adult cortex, and do not show enrichment in certain brain-relevant processes.

## Methods

### Participants

Participants and their parents were recruited through three distinct routes. A total of 57 children who stutter and their parents were recruited during a follow-up visit for the RESTART-randomized trial^36^ in the Netherlands. Participants were included between September 2007 and June 2010 by 24 Speech Language Pathologists in 20 private practice speech clinics throughout the Netherlands. Inclusion criteria were: (1) age between 3;0 and 6;3 years; (2) stuttering was confirmed by a stuttering severity rating on an 8-point scale (at least a score of 2, i.e. ‘mild’) by the parent (3) and the clinician; (4) stuttering frequency was at least 3% syllables stuttered (%SS); and (5) stuttering had been present six months or longer^2^. Exclusion criteria were: (1) diagnosis of an emotional, behavioural, learning or neurological disorder; and (2) lack of proficiency in Dutch for children or parents. For more details, see De Sonneville and others^36^. The medical ethics committee of the Erasmus Medical Center in Rotterdam approved this study (registration number: MEC-2006-349).

All children who were seen for a follow-up visit for the RESTART clinical trial, and their parents, were asked to provide saliva for DNA extraction. For a total of 75 trios, DNA was isolated successfully in high enough concentration and quantity for all three family members. Eighteen trios were excluded from the WES analysis if 1) one or both parents mentioned to have stuttered in the past, or stuttered at the intake of the clinical trial or during follow-up, or 2) > 1 second-degree, > 2 third degree family members, or > 2 second- and third-degree family members were reported to stutter by a parent. The child’s stuttering phenotype (persistent, transient and ambiguous stuttering) was based on parent and teacher ratings (same 8-point scale as described above), and trained observer ratings on the Stuttering Severity Instrument (SSI fourth edition)^37^. Stuttering was categorized as persistent if SSI score ≥ 11, or if SSI score was 9 or 10 and parents or clinician reported presence of stuttering. Stuttering was classified as transient if the SSI score ≤ 8, and both parents and clinician reported absence of any observed stuttering. Conflicts between SSI scores and parents or clinician reports led to categorization of stuttering as ambiguous.

A total of 16 children who stutter and their parents were included via the MPI Erasmus Genetics of Stuttering (MEGS) Study^38^ (https://www.mpi.nl/genetica-van-stotteren). People who stutter were recruited to participate in the MEGS study through national media campaigns, promotion through newspaper articles, television broadcasts, support organizations and social media, and via invitation through speech therapists. Included children and their parents participated between December 2019 and December 2022. Parents of children who stutter were asked to participate in our genetic analyses if 1) their child was 9-15 years of age, 2) their child stuttered at the time of participation, as determined from answering “yes’” to the question “Did your child stutter in the past 12 months”, 3) their child stuttered for at least four years, based on self-reported age at onset of stuttering, 4) both parents reported to have never stuttered, 5) parents reported maximally one second-degree family member who ever stuttered, and 6) the child did not have a diagnosis for ADHD, anxiety, autism, depression, behavioural issues, intellectual disability or hearing difficulty. All children included via MEGS were considered to stutter persistently. Trios were included in the trio WES analysis if DNA of all three family members was available. The medical ethics committee of the Erasmus Medical Center in Rotterdam approved this study (registration number: MEC-2019-0491).

A total of 12 adults who stutter and their parents were included through the Kassel Stuttering therapy center (KST) in Germany. This is a private practice delivering a highly standardized fluency-shaping based therapy, documenting therapy-related changes by standardized videos taken before and after therapy. In 2016, all previous 1450 participants of KST were invited to participate in genetics research by mail, of whom 2033 responded positively, and of whom 180 sent an intact saliva specimen to the cooperating genetic center in Munich. In 2019, the 187 participants were asked to forward parental information leaflets to their parents, encouraging their parents to participate genetic study. Positive replies were received from 108 parents, of whom 33 provided an intact saliva specimen to the cooperating genetic center in Munich. This effort succeeded in assembling 33 trios, of which 12 were included in the present study. All adults who stutter included through the KST were considered to have persistent stuttering. The medical ethics committee of the University of Goettingen approved this study (registration number 19/2/15).

### Whole exome sequencing and variant calling

Whole exome sequencing was performed at the NGS Core Facility, Helmholtz Zentrum, Munich, Germany. Previously published protocols were implemented during sequencing data acquisition and processing.^39^ In short, exome sequences were enriched using SureSelect60Mbv6 library preparation kit (Agilent Technologies, Santa Clara, CA, USA) and sequenced by the means of 100 bp long paired-end reads, produced by the Illumina NovaSeq6000 sequencer (Illumina, San Diego, CA, USA). In-house developed scripts were used to map the reads to the GRCh37/hg19 reference genome sequence (UCSC Genome Browser build hg19 with masked pseudo-autosomal region PAR1 on chromosome Y and updated GRCh38 mitochondrial sequence) with Burrows-Wheeler Aligner (BWA). Single nucleotide variants (SNVs) and small insertions and deletions (indels) were called with SAMTools. All samples were imported into the variant interpretation platform EVAdb of the Institute of Human Genetics, Technical University of Munich, Munich, Germany (https://github.com/IHG-MRI/EVAdb). The *de novo* status of prioritized variants was visually confirmed in Integrative Genomics Viewer (IGV). In addition, all variants classified as (likely) pathogenic or as variant of interest were validated with Sanger sequencing.

### De novo variant identification, annotation and filtering

*De novo* variants were defined as variants that differed from the DNA sequence in both parental samples. The analysis included only small variants (SNVs and indels) within the coding genomic regions with a minimum of 20x coverage. Of those, only non-synonymous variants (missense, nonsense/stop-gain, stop-loss, splice, and frameshift) were kept for the downstream analysis. Genes listed in the actionable incidental findings of the American College of Medical Genetics and Genomics (ACMG SF v3.1, https://www.ncbi.nlm.nih.gov/clinvar/docs/acmg/) were removed prior to the data filtering and interpretation. Variants were filtered based on gene intolerance parameters obtained from the Genome Aggregation Database (gnomAD, v2.1.1)^40^: probable loss-of-function (pLoF) variants were included if the probability of being loss-of-function-intolerant (pLI) was >0.9 and/or if the loss-of-function observed/expected upper bound fraction (LOEUF) was <0.6; missense variants were included if the z-score for missense constraint was >2.5. In addition, pLoF variants were excluded if they were not located in a major transcript, or if they were located within 50 base pairs from the end of the transcript, unless they affected a known functional protein domain. Splice variants were included only if they affected the main acceptor and donor sites.

*De novo* variants that passed these filtering steps were further annotated using ANNOVAR^41^ (version 2017-07-17) with information on minor allele frequencies from gnomAD, measures of evolutionary constraint (Genomic Evolutionary Rate Profiling (GERP++)) and predictions of functional/pathogenic effects used to predict the impact of missense variants on the protein from Mendelian Clinically Applicable Pathogenicity (M-CAP)^42^, rare exome variant ensemble learner (REVEL)^43^ and PrimateAI^44^. Similar predictions from AlphaMissense^45^ were added from https://alphamissense.hegelab.org^46^. M-CAP and REVEL are ensemble methods, based on scores from a combination of often-used tools such as PolyPhen, SIFT and FATHMM, that were shown to outperform these individual tools. PrimateAI classifies variants based on occurrence in other primate species, and AlphaMissense uses a combination of structural context and evolutionary conservation. Together, these four tools use a wide range of evidence to predict the effects of missense variants. Scores from M-CAP > 0.025, REVEL > 0.5, PrimateAI > 0.8 and AlphaMissense > 0.564 were considered to indicate missense variants with damaging effects on protein function, as recommended^42–45^. In addition, for missense variants, conservation estimates of the amino acids carrying a *de novo* variant were obtained from ConSurf^47^. These conservation estimates are based on evolutionary rates in aligned homolog sequences while considering their phylogenetic relationships. ConSurf scores range from 1 (variable) to 9 (conserved).

Expression levels of the genes carrying *de novo* variants were assessed in the developmental human RNA-sequencing dataset of Brainspan^48^ and the adult brain gene expression data in GTEx^49^. Isoform- and exon-specific expression was considered to make sure that the exons carrying the *de novo* variants showed expression in the developing and/or adult brain.

### Variant classification

First, we assessed whether genes previously implicated in monogenic forms of stuttering in multiplex families (*GNPTAB*^12^*, AP4E1*^13^*, IFNAR1*^14^*, ARMC3*^15^*, ZBTB20*^16^ and *PPID*^17^) and hypothesis-driven case/control follow-ups (*GNPTG, NAGPA*^12,18^) carried a *de novo* variant in any of the probands. Second, we investigated whether any gene, regardless of evidence from prior work, harboured recurrent *de novo* mutations in our cohort (i.e. multiple probands carrying a *de novo* mutations in the same gene). Third, we assessed overlaps of genes carrying *de novo* variants in our cohort with genes previously associated with known monogenic disorders. Our focus here was on neurodevelopmental disorders, given the significant comorbidity of speech and language disorders with neurodevelopmental conditions^34,50^, and given that genes previously identified as causal for speech disorders have often also been associated with monogenic neurodevelopmental disorders^30,33^. Searches in PubMed, the Online Mendelian Inheritance in Man (OMIM) database, denovo-db (v1.6.1)^51^, and VariCarta^52^ (assessed in May 2024) were used to identify phenotypes previously linked to similar variants (rare, highly penetrant, and either pLoF or missense) in genes with *de novo* variants. We then classified the highlighted variants into 1) pathogenic, 2) likely pathogenic, 3) uncertain significance, 4) likely benign, and 5) benign variants, by combining layers of evidence of possible impact of the variant to the protein and the trait according to the commonly accepted five-tier classification system for Mendelian disorders.^53^ Fourth, because our approach for interpreting variants has limited power to detect new gene-disease associations, we similarly evaluated evidence of pathogenicity for variants previously not identified as causal for a monogenic neurodevelopmental disorder. This process allowed us to highlight variants of interest in genes of unknown significance.

### Gene set evaluation

To investigate whether there are convergent biological mechanisms that may explain the trait, we created a gene set associated with monogenic forms of stuttering and performed enrichment analyses in datasets that may inform about specific brain processes. This stuttering-associated gene set consisted of twelve genes: the six genes previously associated with monogenic stuttering through genetic investigations of multiplex families: *GNPTAB*^12^*, AP4E1*^13^*, IFNAR1*^14^*, ARMC3*^15^*, ZBTB20*^16^ and *PPID*^17^, as well as the six genes with *de novo* (likely) pathogenic variants and *de novo* variants of interest newly identified in our trio analyses. *GNPTG* and *NAGPA* were not included here, as these genes had been previously associated with stuttering via a hypothesis-driven approach (i.e. in targeted case/control follow-ups of *GNPTAB* variant findings, based on knowledge of functional pathways), so that we could avoid inappropriately biasing enrichments towards the enzymatic mannose 6-phosphate pathway. For a background gene list, we considered only genes that passed the same filtering criteria that we had used for the *de novo* variants, to make sure the results of the enrichment analyses did not reflect our filtering procedure. All 7313 genes intolerant to pLoF or missense variants (pLI ≥ 0.9, LOEUF ≤ 0.6 or mis_z ≥ 2.5), not reported as genes with actionable incidental findings by the ACMG, and not in the stuttering-associated gene list, were included in our background gene list. Next, we assessed enrichment of biological pathways via three complementary approaches, detailed below.

First, we investigated the expression patterns of the stuttering-associated genes in the developing brain. For this, we used regional bulk gene expression data quantified by RNA sequencing from Brainspan (http://www.brainspan.org/) of a total of 224 samples from 23 human brains collected during different developmental periods (8 weeks post conception up to 12 months of age). Gene expression data, measured as fragments per kilobase per million (FPKM), were log-transformed, and then plotted with the package ggplot2 (version 3.4.4) in R (version 4.0.0). A locally estimated scatterplot smoothing (LOESS) curve visualized gene expression change during development. The Brainspan gene expression data were previously clustered into gene expression modules^31^. In short, co-expression similarity of 14,442 genes with high and variable expression was calculated using weighted correlation network analysis (WGCNA)^54^. A total of 16 co-expression modules were detected using the cutreeDynamic hybrid tree cutting function. Module eigengenes were calculated as the first principal component to summarize the expression pattern of the genes in the modules. Enrichment of stuttering-associated genes in the modules, compared to the background gene list, was investigated with two-sided Fisher exact tests. Gene ontology term enrichment analysis to describe the biological processes represented by the modules was performed with GOrilla^55^.

Second, we investigated whether the stuttering gene set was enriched in specific cortical layers or in the white matter of the adult human brain. We made use of spatial gene expression data of the cortex, specifically the inferior frontal gyrus and the posterior part of the superior temporal sulcus; two cortical brain regions relevant to speech and language^56^. This dataset contains spatial gene expression of 48 brain sections (collected from three donors * two brain regions * two tissue blocks * four sections per block) that was measured with Visium spatial transcriptomic slides for a total of 140,192 spots that each represent 3 to 5 cells, in which clustering analysis distinguished twelve data-driven clusters of spots that were related to cortical layers or the white matter. We investigated whether the set of stuttering-associated genes showed differential expression in any of the clusters representing specific cortical layers or white matter, compared to the background gene set. For this, we downloaded the spatial transcriptomics dataset and cluster assignments from^56^ and normalized the count data using 50 principal components calculated from the top 2000 most variable genes using BayesSpace (version 1.12.0) and Harmony (version 1.2.0) packages in R (version 4.3.1) as was done previously^56^. Next, we pseudobulked the spot-level data for each gene into cluster-level data by averaging the normalized counts for the respective gene across all spots in a given cluster with the summarizeAssayByGroup function of the scuttle R package (version 1.15.4). Then, we log-transformed the averaged counts, and plotted their distributions using the ggplot2 R package. The spatial brain expression pattern of the stuttering gene set was compared to that of the background gene set across the clusters representing cortical layers.

Third, we investigated whether common variants (single nucleotide polymorphisms; SNPs) in and near the stuttering-associated genes affect the strength of white matter connections in the adult human brain. For this, we made use of results of multivariate genome-wide association studies (GWAS) on white matter connectivity^57^. GWAS results for node- and edge-level measures of white matter connectivity were downloaded from the NHGRI-EBI GWAS Catalog^58^ (study ID GCST90165317 and GCST90165318, downloaded June 2024). SNP-level p-values were converted to gene-based p-values using Multi-marker Analysis of GenoMic Annotation (MAGMA version 1.10)^59^: SNPs were linked to a gene if they were located within the gene body or an 15kb upstream gene window. Next, a gene-set enrichment analysis was performed that tests whether the gene-based p-values of stuttering-associated genes are lower than those of genes in the background gene list, while correcting for gene size, the level of linkage disequilibrium between SNPs in the gene, and the inverse of the minor allele count of the SNPs in the gene.

## Results

A total of 85 parent-offspring trios was included to identify potential pathogenic variants that may explain the stuttering in the children (Table 1; Supplemental Table 1). High-quality WES data were generated from 84 trios and one parent-child duo; the latter because for one father high-quality WES data could not be generated from the available DNA sample. We searched for *de novo* variants in the sequencing data of the 84 complete trios by excluding all variants present in any of the unaffected parents. Across the whole cohort, a total of 383 *de novo* non-synonymous variants were called. No *de novo* variants were identified in any of the genes previously associated with monogenic forms of stuttering. In addition, no gene was identified with recurrent *de novo* variants, regardless of prior evidence about relevance for stuttering.

**Table 1:** overview of participants.

|  | RESTART | MEGS | KST cohort | Total |
| --- | --- | --- | --- | --- |
| **N (% males)** | 57 (77%) | 16 (81%) | 12 (67%) | 85 (75%) |
| **N persistent stuttering (% males)** | 19 (100%) | 16 (81%) | 12 (67%) | 47 (85%) |
| **N transient stuttering (% males)** | 30 (57%) | 0 | 0 | 30 (57%) |
| **N ambiguous stuttering (% males)** | 8 (88%) | 0 | 0 | 8 (88%) |
| **Mean age in years (range)\*** | 10.6 (8-13) | 10.8 (9-14) | 33.9 (26-49) | 13.2 (8-49) |

\*Age information is missing for three participants of the KST cohort.

### Pathogenic and likely pathogenic *de novo* variants identified in probands who stutter

We identified three *de novo* probable-loss-of-function (pLoF) variants in constrained genes (Table 2). One of the pLoF variants was found in a gene previously linked to a neurodevelopmental disorder: the stop-gain in *SPTBN1* (c.A520T; p.R174X) in proband MEGS_14 (Figure 1). Missense and pLoF variants in *SPTBN1* were recently implicated in a neurodevelopmental disorder characterized by intellectual disability, language and motor delays, autistic features and seizures^60,61^. This variant was therefore classified as pathogenic. Still, replication is required to confirm stuttering is a feature of the monogenic neurodevelopmental disorder associated with mutations in *SPTBN1*. The two other pLoF variants are located in *FLT3* and *IREB2*; two genes not associated with a neurodevelopmental disorder. Because pLoF variants in these genes are highly uncommon, we classified these variants as variants of interest, even though additional evidence is required to verify a causal relation between the two genes and stuttering or other neurodevelopmental disorders.

**Figure 1:**
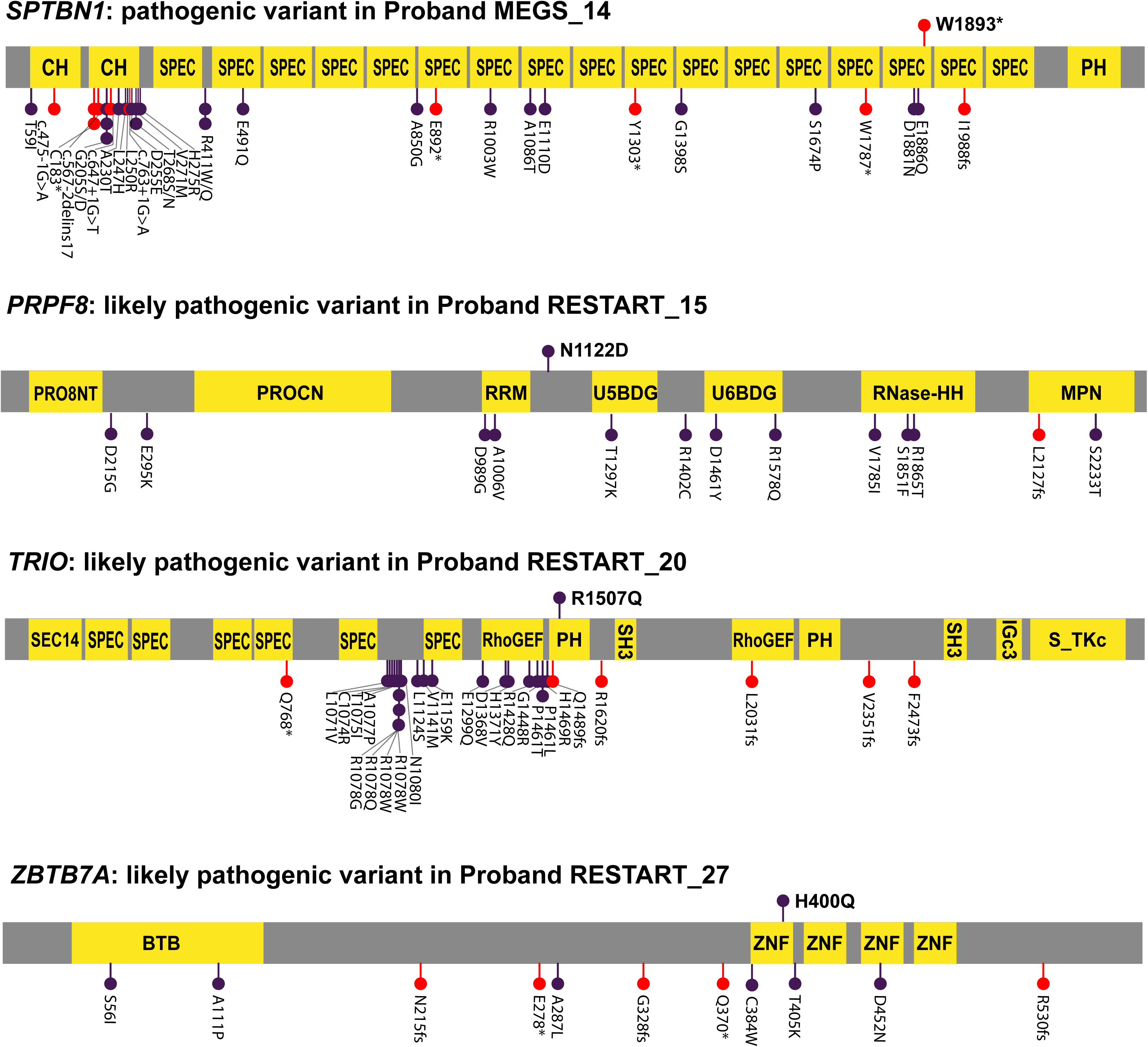
Locations of identified (likely) pathogenic variants in stuttering and published pathogenic neurodevelopmental disorder variants in the same genes. Pathogenic and likely pathogenic variants identified in this study are visualized above the linear protein schematics. The variants previously published as causal for monogenic neurodevelopmental disorders related to *SPTBN1*^60,61^, *PRPF8*^62^*, TRIO*^64,65^ and *ZBTB7A*^66,67^ are visualized below the schematics. Missense variants are indicated in purple and pLoF variants in red. Protein domains are represented with yellow squares: CH: calponin homology domain; SPEC: spectrin repeats; PH: Pleckstrin homology domain; PRO8NT: PrP8 N-terminal domain; PROCN: central domain in pre-mRNA splicing factors of PRO8 family; RRM: RNA recognition motif; U5/6BDG: U5/6-snRNA binding site; RNase-HH: RNase-H homology domain; SEC14: protein structural domain that binds small lipophilic molecules; RhoGEF: guanine nucleotide exchange factor; SH3: Src homology 3; S_TKc: Serine/Threonine protein kinases, catalytic domain; BTB: Broad-Complex, Tramtrack and Bric a brac; ZNF: Zinc finger.

**Table 2:** D*e novo* frameshift and nonsense variants identified in people who stutter.

| Proband | St | Gene | Variant type | Isoform | cDNA change | protein change | pLI | LOEU F | MA F | NDD gene | Classification criteria* | Classification* |
| --- | --- | --- | --- | --- | --- | --- | --- | --- | --- | --- | --- | --- |
| **RESTART_11** | P | *FLT3* | Stop-gain | NM_004119 | c.A520T | p.R174X | 0.61 | 0.35 | 0 | No | - | Variant of interest in GUS |
| **MEGS_14** | P | *SPTBN1* | Stop-gain | NM_003128 | c.G5678A | p.W1893X | 1.00 | 0.09 | 0 | Yes | PVS1, PS2, PM2 | Pathogenic |
| **RESTART_8** | A | *IREB2* | Frameshift | NM_004136 | c.619delC | p.P207QfsTer9 | 1.00 | 0.22 | 0 | No | - | Variant of interest in GUS |

\*Classification criteria and classification based on ACMG guidelines<sup>53</sup>. St: stuttering, classified as P: Persistent and A: ambiguous. pLI; probability of being loss-of-function intolerant. LOEUF: loss-of-function observed/expected upper bound fraction. MAF: minor allele frequency. NDD: neurodevelopmental disorder. GUS: gene of unknown significance.

We identified twelve *de novo* missense variants that passed our filtering criteria (Table 3). A total of six *de novo* missense variants were located in genes previously identified as causal for a neurodevelopmental disorder, of which three variants in *PRPF8*, *TRIO* and *ZBTB7A* were classified as likely pathogenic (Figure 1). In addition to passing the filtering criteria (the variants are *de novo* and are located in a constrained gene), all three variants are (i) absent from or seen only once in GnomAD, (ii) located in a conserved region of the gene as evident by ConSurf and GERP++, and (iii) considered damaging by all *in silico* tools used to predict pathogenicity of missense variants. Proband RESTART_15 carries a likely pathogenic missense variant in *PRPF9*. This gene encodes a scaffolding component of a spliceosome complex that splices pre-mRNA into mRNA by removing introns. Recently, missense and LoF variants located throughout the protein have been identified as the cause of a neurodevelopmental condition involving developmental delay and autism^62^. In addition, heterozygous missense variants in the C-terminal MPN-domain cause autosomal dominant retinitis pigmentosa^63^. *TRIO* encodes a Rho guanine nucleotide exchange factor (RhoGEF) that has previously been identified as causal for neurodevelopmental disorders, with domain-specific symptoms^64,65^. Gain-of-function missense variants in and near the spectrin domains are associated with severe developmental delay, speech and language delay and macrocephaly, while loss-of-function variants and missense variants in the RhoGEF domain show milder developmental delay, speech and language delay, and microcephaly. The p.R1507Q *TRIO* missense variant in proband RESTART_20 is located slightly upstream of the RhoGEF domain, in the PH domain that assists and regulates the activity of the RhoGEF domain.

**Table 3:** D*e novo* missense variants identified in people who stutter.

| Proband | St | Gene | Isoform | cDNA change | protein change | mis_Z | MAF | M-CAP | REVEL | Primate AI | Alpha Missense | GERP ++ | Con Surf | NDD gene | Classification criteria* | Classification* |
| --- | --- | --- | --- | --- | --- | --- | --- | --- | --- | --- | --- | --- | --- | --- | --- | --- |
| RESTART_15 | P | *PRPF8* | NM_006445 | c.A3364G | p.N1122D | 8.3 | 0 | D | D | D | D | 5.31 | 9 | Yes | PS2, PM2, PP2, PP3 | Likely pathogenic |
| RESTART_47 | P | *CHD4* | NM_001273 | c.A2477G | p.N826S | 6.3 | 8.0x10<sup>-6</sup> | D | T | D | T | 4.9 | 5 | Yes | PS2, PP2, PP3, BP4 | VUS |
| RESTART_56 | P | *UNC13A* | NM_001080421 | c.T224C | p.V75A | 5.6 | 0 | D | D | T | A | 4.98 | 7 | No | - | - |
| KST_2 | P | *EIF2AK4* | NM_001013703 | c.G3040A | p.V1014M | 2.7 | 0 | T | T | T | T | 5.82 | 8 | No | - | - |
| KST_4 | P | *GIT1* | NM_001085454 | c.G2135T | p.R712L | 3.1 | 0 | D | T | T | A | 4.65 | 9 | No | - | - |
| KST_7 | P | *ADGRB1 / BAI1* | NM_001702 | c.G1441T | p.A481S | 4.5 | 4.2x10<sup>-6</sup> | T | T | T | T | 3.55 | 4 | No | - | - |
| KST_10 | P | *TUT4 / ZCCHC11* | NM_015269 | c.A1442T | p.D481V | 2.8 | 0 | D | T | T | A | 5.51 | 7 | No | - | - |
| RESTART_18 | T | *PLXNA1* | NM_032242 | c.G3182C | p.S1061T | 3.4 | 0 | D | D | T | A | 3.84 | 7 | Yes | PS2, PM2, PP2, PP3, BP4 | VUS |
| RESTART_20 | T | *TRIO* | NM_007118 | c.G4520A | p.R1507Q | 5.3 | 3.2x10<sup>-5</sup> | D | D | D | D | 5.39 | 9 | Yes | PS2, PP2, PP3 | Likely pathogenic |
| RESTART_22 | T | *NCDN* | NM_001014839 | c.C2095T | p.R699W | 3.8 | 0 | D | T | T | T | 5.13 | 1 | Yes | PS2, PM2, PP2, BP4 | VUS |
| RESTART_27 | T | *ZBTB7A* | NM_015898 | c.C1200G | p.H400Q | 4.0 | 0 | D | D | D | D | 4 | 9 | Yes | PS2, PM2, PP2, PP3 | Likely pathogenic |

\*Classification criteria and classification based on ACMG guidelines<sup>53</sup>. St: stuttering, classified as P: Persistent and T: transient. MAF: minor allele frequency. Scores of M-CAP, REVEL, PrimateAI and AlphaMissense were interpreted as D: deleterious, T: tolerated, and A: ambiguous. NDD: neurodevelopmental disorder. GUS: gene of unknown significance. VUS: variant of unknown significance

Variants in *ZBTB7A* cause a neurodevelopmental disorder characterized by intellectual disability, macrocephaly, and overgrowth of adenoid tissue^66^. The p.H400Q variant in proband RESTART_27 is located in the first zinc finger domain, which is also the location of two previously described missense variants^66,67^. Yet, none of the individuals previously described with a pathogenic mutation in *PRPF8*, *TRIO*, or *ZBTB7A* have been described to stutter. Similarly, lack of evidence suggests that probands RESTART_15, RESTART_20 and RESTART_27 do not show symptoms of the severe neurodevelopmental problems typically associated with pathogenic mutations in *PRPF8*, *TRIO*, and *ZBTB7A.* Despite cumulative evidence that the missense variants affect the proteins, additional support such as identifying the same variants in other individuals with a neurodevelopmental disorder or functional validation of an effect on the protein would be needed to fully prove that these variants are pathogenic. In addition, replication in a person who stutters is required to confirm that stuttering is a feature of the monogenic neurodevelopmental disorders associated with mutations in *PRPF8*, *TRIO*, and *ZBTB7A*.

The missense variants in *CHD4*, *PLXNA1,* and *NCDN* were classified as variant of unknown significance, because computational evidence suggested a less deleterious or tolerated effect of the variants on the proteins. Even though the p.N826S variant in *CHD4* in proband RESTART_47 is located in the ATPase domain, where multiple disease-causing variants are aggregated^68^, the asparagine at position 826 is not as highly conserved as the amino acids mutated in patients with CHD4-related syndrome (ConSurf score of 5 [average], compared to 7-9 [conserved]). It is therefore unlikely that this missense variant has a major effect on the functioning of the CHD4 protein. However, *in silico* prediction tools of effects of variants on proteins currently cannot fully capture true effects. Additional evidence would therefore be required to conclusively classify these variants as pathogenic or benign according to standard criteria.

### Gene-set analyses to investigate biological pathways involved in monogenic stuttering

We investigated whether genes associated with monogenic forms of stuttering share roles in brain-relevant cell types and (developmental) processes. To do so, we tested for enrichment of all genes so far linked with monogenic stuttering in relevant datasets. In the stuttering gene set, we included the genes identified in the current trio analysis with (likely) pathogenic *de novo* variants (*SPTBN1*, *PRPF8*, *TRIO* and *ZBTB7A*) and with variants of interest (*FLT3* and *IREB2*), as well as the six genes associated with stuttering through previous family-based rare variant investigations (*GNPTAB*, *AP4E1*, *IFNAR1*, *ARMC3*, *ZBTB20* and *PPID*). First, we investigated whether these twelve stuttering-associated genes show similar gene expression patterns in the developing brain. Similar expression during brain development is observed for genes implicated in a number of neurodevelopmental disorders including childhood apraxia of speech^31,35^ and autism spectrum disorder^69^. In contrast, these twelve stuttering-associated genes show very dissimilar expression patterns during brain development (Figure 2A). To investigate whether a particular expression pattern is overrepresented, we made use of gene co-expression modules of the same dataset. Gene expression modules consist of genes co-expressed in certain regions of the brain during development (Supplemental Figure 1), and are enriched for brain-relevant developmental processes (Supplemental Table 2). Nine genes associated with monogenic forms of stuttering were assigned to a module. They were present in six of the sixteen modules, representing processes including synapse organization, transcription factor activity and chromatin organization. A maximum of two stuttering-associated genes were assigned to the same module, and none of the modules were enriched for stuttering-associated genes, confirming limited co-expression of the genes. *PRPF8* and *TRIO* were present in the module previously found enriched for genes linked to childhood apraxia of speech. Yet, none of the modules showed an enrichment of the stuttering-associated gene set.

**Figure 2:**
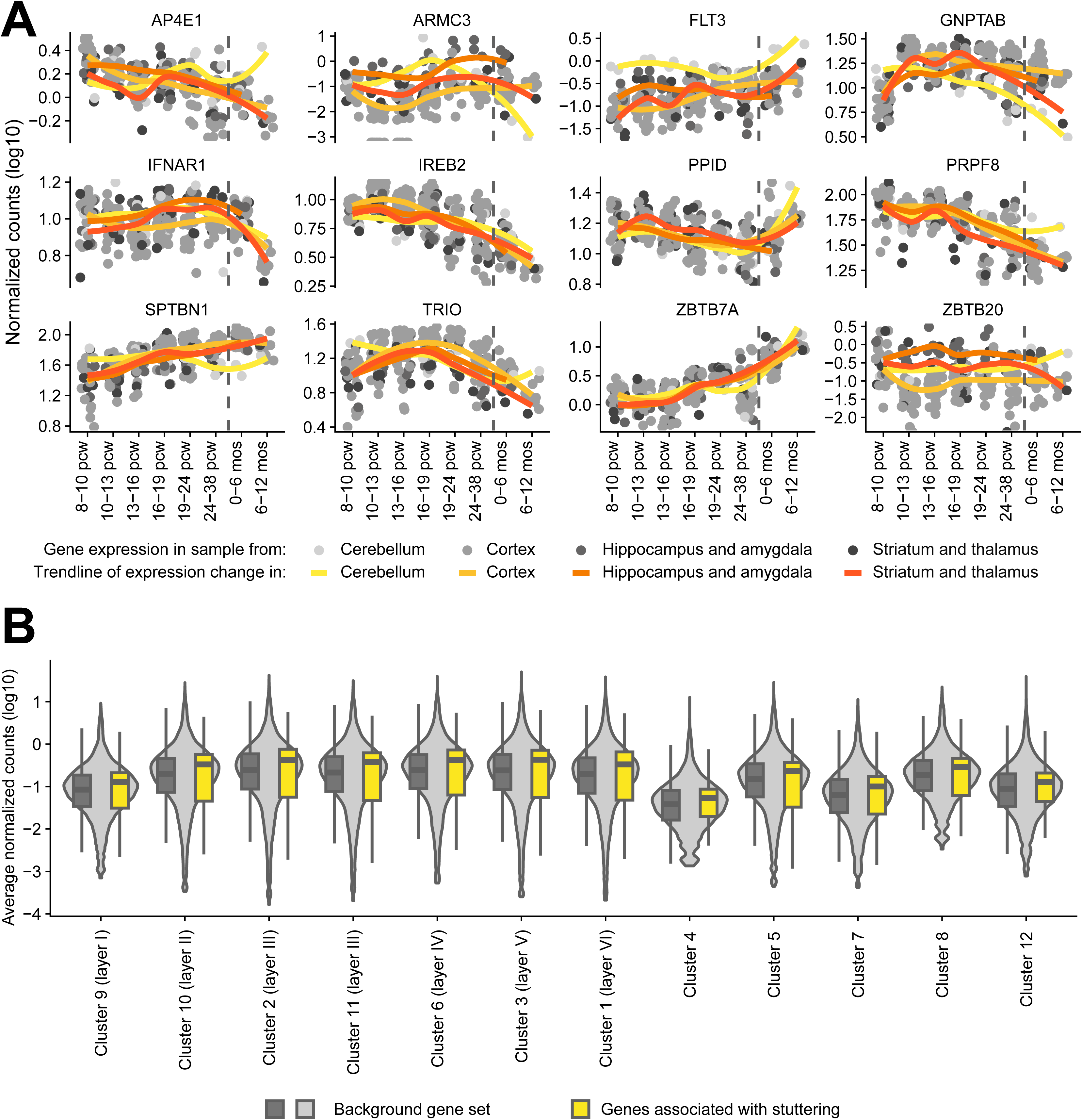
Neural gene expression patterns of genes associated with monogenic forms of stuttering. **A.** Developmental brain expression pattern of the twelve stuttering-associated genes across eight developmental periods spanning from eight post conception weeks (pcw) to ten months (mos) of age. Grey circles depict expression levels in individual brain samples collected from the cerebellum, cortex, hippocampus, amygdala, striatum and thalamus. The trendlines in yellow-orange (estimated with locally estimated scatterplot smoothing) visualize the overall pattern of gene expression change over time in the different regions of the brain. The vertical dashed lines represent time of birth. **B.** Gene expression levels of these stuttering-associated genes and the background gene set in spatial gene expression data of the adult human cortex. Spatial gene expression data of 48 human cortex tissue sections were clustered into 12 data-driven clusters, of which seven represent cortical layers and five were located in the white matter^56^. Violin plots and grey box plots show the distribution of gene expression levels of the background gene set in the 12 clusters. Yellow box plots show the gene expression levels of the stuttering-associated genes. Box plots show median and first and third quartiles, with whiskers extending to 1.5 times the interquartile range.

Second, we investigated whether genes linked to monogenic forms of stuttering show specific spatial expression patterns in the adult human brain. For this, we made use of spatial transcriptomics data of the human cortex^56^. Spatial transcriptomics is a technique that measures gene expression for many thousands of transcripts in a tissue section, across several thousands of locations (spots), in this case each representing three to five cells. Data-driven clustering of this dataset identified seven clusters that recapture the laminar structure of the cerebral cortex, and five clusters that were located in the white matter. The twelve genes associated with monogenic stuttering show gene expression levels that are very similar to the background gene set in each of the clusters, and do not highlight a certain cortical layer or white matter cluster (Figure 2).

Third, we investigated whether the genes so far associated with monogenic forms of stuttering play a role in human brain white matter connectivity. Previously, several lines of evidence indicated a role for reduced white matter connectivity in stuttering. Because our gene expression analyses failed to identify overlapping expression patterns or biological functions between the stuttering-associated genes, we also applied a more direct approach to explore the relation between the stuttering-associated genes and white matter connectivity. For this, we made use of results of a recent multivariate GWAS for white matter connectivity of the human brain, that used brain imaging data of 30,810 individuals of the UK Biobank^57^. The authors of that study used fiber tractography of diffusion tensor imaging data, to derive two measures of connectivity, where the nodes captured the sum of the connectivity of each of the 90 brain regions investigated, and the edges captured the connectivity between 947 pairs of brain regions. We converted SNP-based p-values from the two multivariate genome-wide analyses into gene-based p-values. Three of the twelve stuttering-associated genes showed low gene-based p-values for one or both measures of white matter connectivity (Table 4). We next tested whether the set of stuttering-associated genes was enriched for low p-values. For both the node-level connectivity (beta=0.17, se=0.33, p=0.31) and edge-level connectivity (beta=-0.02, se=0.35, p=0.53) GWASs, there was no enrichment of low gene-based p-values in our stuttering gene set. So, while variation in some stuttering-associated genes may be associated with variability in white matter connectivity, alterations of the latter may not be a common mechanism that is shared across genes implicated in monogenic forms of stuttering.

**Table 4.** Gene-based p-values of stuttering-associated genes in human brain white matter connectivity GWAS in 30,810 individuals.

| Stuttering-associated gene | Number of SNPs assigned to gene | Gene-based p-value for node-level connectivity | Gene-based p-value for edge-level connectivity |
| --- | --- | --- | --- |
| *GNPTAB* | 252 | 0.012 | 0.23 |
| *AP4E1* | 307 | 0.011 | 0.014 |
| *IFNAR1* | 196 | 0.69 | 0.33 |
| *ARMC3* | 338 | 0.24 | 0.12 |
| *ZBTB20* | 1641 | 0.049 | **0.0014** |
| *PPID* | 44 | 0.027 | 0.42 |
| *SPTBN1* | 761 | **5.49x10<sup>-4</sup>** | **5.84x10<sup>-4</sup>** |
| *PRPF8* | 197 | 0.54 | 0.60 |
| *TRIO* | 1070 | 0.042 | 0.39 |
| *ZBTB7A* | 101 | **4.03x10<sup>-5</sup>** | **9.38x10<sup>-5</sup>** |
| *FLT3* | 520 | 0.62 | 0.52 |
| *IREB2* | 188 | 0.53 | 0.60 |

Gene-based p-values were calculated with MAGMA from two multivariate GWAS analyses of 90 node-level and of 851 edge-level connectivity measures of the human brain<sup>57</sup>. P-values are marked bold if significant after Bonferroni-correction for 24 tests (p-value threshold is 0.0021).

## Discussion

Here, we used whole exome sequencing of 85 children who stutter to identify genes potentially involved in monogenic forms of stuttering. By including parents who had never stuttered, our trio study design enabled us to identify and focus our analyses on *de novo* variants in the children who stutter. To our knowledge, this is the first time *de novo* variants have been implicated in stuttering, as previous studies in this area have focused on families in which multiple relatives stutter^12–17^. We identified a *de novo* stop-gain variant in *SPTBN1* that could be classified as pathogenic, and missense variants in *PRPF8*, *TRIO* and *ZBTB7A* that could be classified as likely pathogenic. In addition, likely damaging *de novo* variants in genes not previously implicated in neurodevelopmental disorders highlighted two genes of interest for stuttering: *FLT3* and *IREB2*. Our yield of four (likely) pathogenic variants in 84 trios indicates that *de novo* variants are not a major cause of stuttering, and is notably lower than yields previously found for childhood apraxia of speech (36 in 122 probands across three studies)^30^ and speech delay (three in 23 probands)^33^. Still, we show that rare *de novo* variants might account for a subset of cases and so should not be neglected as a possible cause for stuttering.

Our study additionally represents the first rare-variant analysis to include not only persistent cases but also individuals with transient developmental stuttering. All previous rare-variant investigations of stuttering focused on individuals with persistent developmental stuttering^12–17^. Surprisingly, our yield of (likely) pathogenic variants did not differ between the groups with persistent (two in 47 probands) and transient (two in 30 probands) stuttering. Moreover, beyond these (likely) pathogenic variants, our study highlighted two further genes of interest: in a proband with persistent stuttering, and a proband whose stuttering was classified as ambiguous. Little is known about differences and similarities in the genetic foundations of transient and persistent stuttering. A twin study in 12,892 children that distinguished transient and persistent stuttering showed very similar heritability estimates (h2=67% and 60%, respectively), and also identified multiple occurrences of transient and persistent stuttering within a twin pair^7^. In addition, an investigation of inheritance patterns in the extended families of 66 children who stutter found that transient and persistent stuttering have a shared genetic basis, and that persistent stuttering may at least in part be caused by additional (genetic) factors^70^.

Another innovative aspect of our findings is the novel evidence of a direct genetic link between stuttering and other neurodevelopmental disorders. Even though the genes *AP4E1* and *ZBTB20* have previously been linked to stuttering^13,16^ and separately to neurodevelopmental disorders, the mode of inheritance (i.e. dominance/recessivity) do not overlap. Heterozygous missense variants in *ZBTB20* cause Primrose syndrome^29^, while the gene was associated with stuttering through a recessive mode of inheritance^16^. Similarly, biallelic loss-of-function variants in *AP4E1* cause spastic paraplegia^28^, while a haplotype of two missense variants as well as heterozygous variants have been associated with stuttering^13^. According to prior literature on the genes implicated by our *de novo* analyses, none of the patients with neurodevelopmental disorders caused by mutations in *SPTBN1*^60,61^, *PRPF8*^62^, *TRIO*^64,65^, and *ZBTB7A*^66^ have been described to stutter. Stuttering may be an uncommon feature of these neurodevelopmental disorders, but an alternative explanation is that stuttering diagnoses may have escaped detection, because stuttering is often not registered well in electronic medical records^50^. The latter is supported by the increased prevalence of developmental conditions including intellectual disability, learning disability, seizures and ADHD in children who stutter^71^. Interestingly, a likely pathogenic missense variant in *SPTBN1* has been described in a proband with speech delay^33^, and a missense variant of unknown significance in *TRIO* in a proband with childhood apraxia of speech^31^. In addition, pathogenic variants in *SPTBN1* have been associated with aphasia^50^. Lastly, more general terms for speech difficulties such as delayed speech, expressive and/or receptive language difficulties and absence of speech have been registered for many patients with neurodevelopmental disorders related to *SPTBN1*^60,61^, *TRIO*^64,65^, and *ZBTB7A*^66^, although not for *PRPF8*^62^. Detailed speech and language analysis in people with mutations in neurodevelopmental disorder genes including *KAT6A*^72^*, SETBP1*^73^, and *BRPF1*^74^, that were performed after identification of a (likely) pathogenic variant in these genes in an individual with childhood apraxia of speech, revealed widespread speech and language difficulties. Such phenotypic assessments highlight that speech and language difficulties are usually not systematically investigated. Identification of rare pathogenic variants that cause stuttering and other speech disorders may thus point towards neurodevelopmental disorders in which speech difficulties are a central feature. It is now important to further prove a role for *SPTBN1*, *PRPF8*, *TRIO* and *ZBTB7A*, *FLT3* and *IREB2* in stuttering by identifying recurrent mutations in other people who stutter, or through extensive assessments of the speech phenotypes in people with a mutation in any of these genes.

Several lines of evidence from genetic and brain imaging studies suggest the involvement of altered white matter in stuttering. First, different transgenic mouse models carrying putative pathogenic variants of *GNPTAB* or *PPID* both showed white-matter features that distinguished the knock-in animals from wild-type animals (although the nature of these features differed)^17,25^. Second, *SPTBN1* (newly implicated in the present study) encodes a cytoskeletal protein important for axonal formation and function^75^. Third, several brain imaging studies in adults and children who stutter have reported decreased white matter integrity, most commonly along parts of the left arcuate fasciculus and/or superior longitudinal fasciculus, white-matter tracts which connect parts of the frontal cortex with cortical areas in the parietal and temporal lobes^76^. We therefore investigated whether the genes thus far linked to monogenic forms of stuttering show enrichment of common variants involved in white-matter connectivity. A few of the stuttering-associated genes: *SPTBN1, ZBTB20*, and *ZBTB7A*, showed significant gene-based association with measures of white-matter connectivity. Yet, the full set of the twelve stuttering-associated genes that we investigated here did not show an enrichment of genetic associations with white matter connectivity as derived from GWAS data.

Genes causally implicated in neurodevelopmental disorders with similar features often show overlapping gene expression patterns in the brain and overlaps in functional pathways. For example, the majority of genes thus far implicated in childhood apraxia of speech regulate gene expression through transcription factor activity or chromatin remodeling, and are highly expressed at early stages during brain development^31,32^. However, the genes thus far linked to monogenic forms of stuttering, through previous family-based investigations and the current trio analysis, do not converge onto one or a few shared processes. The developmental brain expression data and analysis method that previously showed overlaps among genes causal for childhood apraxia of speech^31^ and among genes implicated in autism spectrum disorder^69^, here found no significant overlaps in expression patterns among genes associated with monogenic forms of stuttering. A similar lack of convergence was seen when using spatial gene expression data of the human adult brain. Our results may indicate that stuttering can result from differences in a broad range of biological processes and brain regions/cell-types. Alternatively, the current analyses may overlook the biological processes, brain regions, or developmental periods involved, either because they were undersampled or because the bulk and spatial gene expression data did not have the resolution to detect a signal. Other datasets or additional implicated genes may be required to identify convergent processes, if these exist.

Our study has several limitations. First, the exploration of *de novo* variants may overlook inherited causal variants with reduced penetrance, regulatory variants not located in the exons, and repeat expansions. Even though we selected probands with limited stuttering reported in family members, thereby optimizing our study design for the identification of *de novo* variants with high effect sizes, variants with low penetrance may explain cases who did report a few family members who stutter, or who failed to report transient stuttering of family members. Second, our filtering to include and exclude variants as likely pathogenic strongly depends on prediction tools that inform about how damaging a variant may be to a protein. Even though we combined evidence from four prediction tools that are based on different types of information and thus may be seen as supplementary layers of evidence, over- or underestimation of the effects of variants may have led us to wrongly include or exclude variants. Recurrent findings or functional testing (*in vitro* or *in vivo*) may provide final evidence for a pathogenic or benign role of variants classified as likely pathogenic or VUS. Third, our yield cannot inform about how prevalent monogenic forms of stuttering are, because we only investigated *de novo* variants and thus do not have information about rare inherited causal variants. Fourth, we currently cannot verify that the (likely) pathogenic variants cause the stuttering in the probands. Even after our careful and strict variant filtering and classification process, we can only judge whether the variants may be (likely) pathogenic for the neurodevelopmental traits previously associated with the gene. Identification of additional pathogenic variants in the same genes in people who stutter, or extensive phenotypic analysis of the speech of people with a mutation in any of these genes, is required to prove that these (likely) pathogenic variants cause stuttering.

In conclusion, by analyzing genome sequencing data of 85 individuals with persistent, transient or ambiguous stuttering and parents who do not stutter, we identified rare *de novo* variants of which four were classified as (likely) pathogenic and two highlighted genes of interest. We linked stuttering to genes causal for other monogenic neurodevelopmental disorders with and without speech problems. Extensive analysis of two brain gene expression datasets and a neuroimaging GWAS dataset indicates that monogenic forms of stuttering are likely to involve heterogenous biological pathways, rather than a shared mechanistic basis.

## Data Availability

All data produced in the present study are available upon reasonable request to the authors

## Acknowledgements

EE, AV and SEF are financially supported by the Max Planck Society. EE is also supported by a Veni grant of the Dutch Research Council (NWO; VI.Veni.202.072).

RESTART study: We gratefully acknowledge all participants, their parents and speech therapists for their participation in the RESTART trial. In addition, we acknowledge the contributions of Caroline de Sonneville-Koedoot, Nina Koemans and Sophie Jacobs to the collection of the follow-up data; of Sarah Graham to the collection of DNA; and of Esther Bunschoten, Chris Struiksma, Conor Jansen en Nienke van den Broek to the rating of stuttering severity from the speech recordings.

MEGS: We are grateful to the children and their parents who participated in the MEGS trio study. We also thank Loes van den Heuvel, Cansel Sert, and Anke Verhulst for their assistance regarding data collection.

KST cohort: The authors are thankful to the KST and their staff for long term cooperation, to the recently dissolved Department of Clinical Neurophysiology, University Medical Center Göttingen, for administrative support including most postal service charges. They acknowledge receiving a seed grant from the Leibniz Wissenschaftscampus primate cognition to M.S. and to Prof. Dr. Nivedita Mani, Georg-Elias-Müller Institute for Psychology, Göttingen Sommer/Mani DM 22-606) for assembling a video-archive allowing detailed characterization of the phenotype of 187 participants for which a saliva sample is available.

---

**Supplemental Figure 1:**
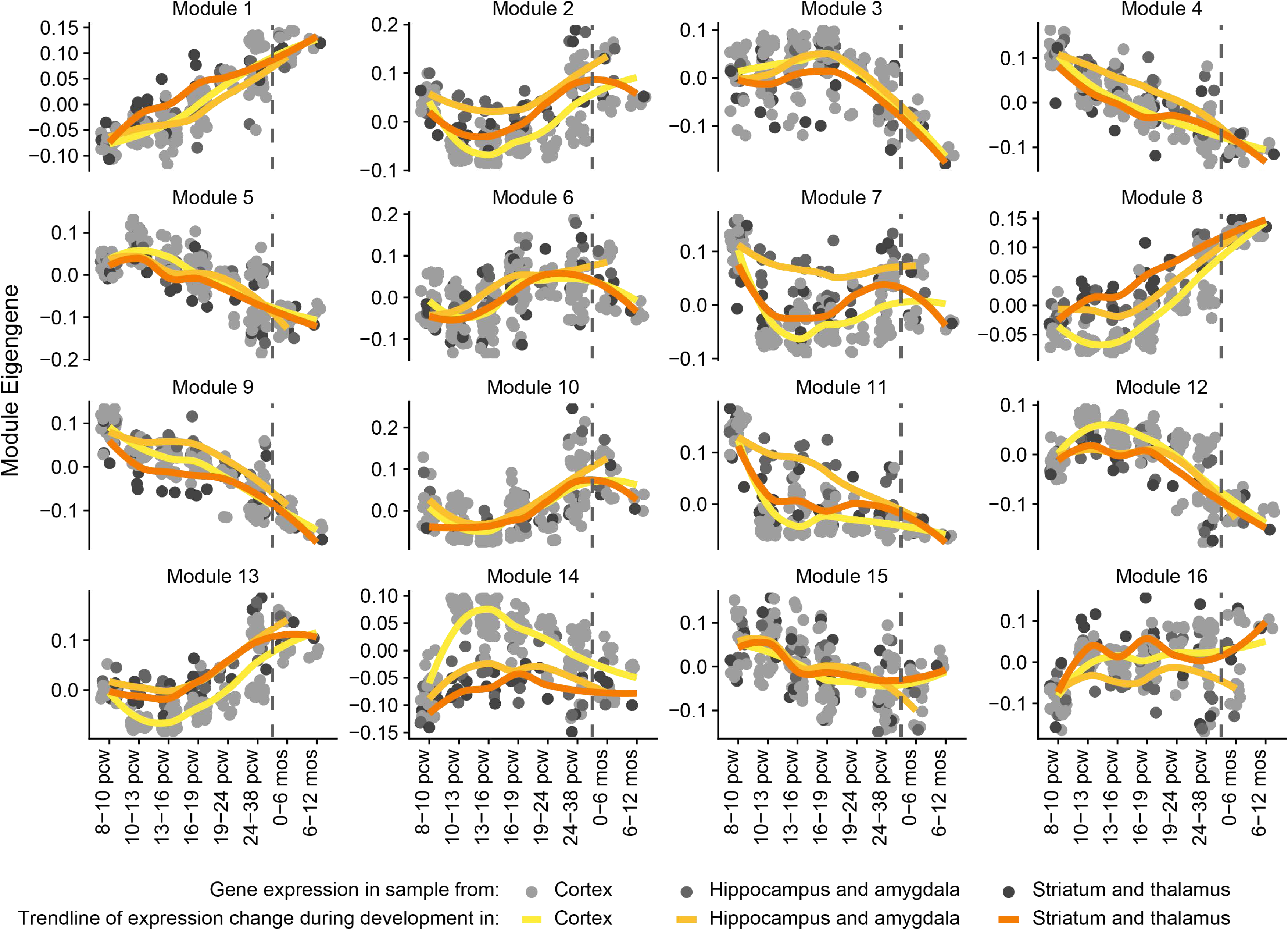
Expression pattern of gene expression modules during brain development. Module Eigengenes represent the overlapping expression pattern of all genes represented by the module. Each dot represents a brain sample, the yellow lines are the loess curve fitted through the data points. The vertical dashed lines represent time of birth. Pcw: post conception week.

## Supplemental Tables

**Supplemental Table 1:** overview of participants.

| Proband | St | Sex | Age@ at participation | Age@ at onset | Self-report speech fluency<sup>#</sup> | Parent-report speech fluency<sup>#</sup> | Therapist-report speech fluency | SSI | Trialled therapies | Relatives who stutter(ed) | Other diagnoses |
| --- | --- | --- | --- | --- | --- | --- | --- | --- | --- | --- | --- |
| RESTART_1 | T | Male | 11-15 | 1-5 | Mild / mild to moderate | Borderline | - | 2 | Lidcombe program | No | Dyslexia |
| RESTART_2 | P | Male | 11-15 | 1-5 | Mild | Mild | Fluent | 11 | RESTART-DCM | No | Dyslexia |
| RESTART_3 | T | Female | 11-15 | 1-5 | Fluent | Fluent | Fluent | 6 | RESTART-DCM | 2<sup>nd</sup> degree relative | No |
| RESTART_4 | T | Male | 11-15 | 1-5 | Fluent | Fluent | Fluent | 2 | Lidcombe program | 4<sup>th</sup> degree relative | No |
| RESTART_5 | T | Male | 11-15 | 1-5 | Fluent | Fluent | Fluent | 6 | Lidcombe program | No | Dyslexia |
| RESTART_6 | P | Male | 11-15 | 1-5 | Borderline | Fluent | Fluent | 13 | Lidcombe program | No | No |
| RESTART_7 | P | Male | 11-15 | 1-5 | Mild to moderate | Mild | Mild | 12 | Lidcombe program | No | Dyslexia |
| RESTART_8 | A | Male | 11-15 | 1-5 | Mild | Mild | Fluent | 0 | Lidcombe program | No | Behaviour problems, social/emotional problems, autism |
| RESTART_9 | A | Female | 11-15 | 1-5 | Borderline | Fluent | Fluent / borderline | 12 | Lidcombe program | No | No |
| RESTART_10 | T | Male | 11-15 | 1-5 | Borderline | Fluent | Fluent | - | Lidcombe program | No | No |
| RESTART_11 | P | Male | 11-15 | 6-10 | Mild to moderate | Mild to moderate | Mild to moderate | 25 | RESTART-DCM | No | Social/emotional problems, hypermobility |
| RESTART_12 | T | Male | 11-15 | 1-5 | Fluent | Fluent | Mild | 2 | Lidcombe program | No | Add/adhd |
| RESTART_13 | T | Female | 11-15 | 1-5 | Borderline | Fluent | Fluent / borderline | 2 | Lidcombe program | 2<sup>nd</sup> and 4<sup>th</sup> degree relative | Social/emotional problems |
| RESTART_14 | A | Male | 6-10 | 1-5 | Mild to moderate | Fluent | Fluent / borderline | - | RESTART-DCM | No | No |
| RESTART_15 | P | Male | 11-15 | 1-5 | Moderate | Moderate | Mild | - | Lidcombe program | 2<sup>nd</sup> degree relative | No |
| RESTART_16 | T | Female | 11-15 | 1-5 | Borderline | Fluent | Fluent | - | Lidcombe program | No | No |
| RESTART_17 | T | Female | 11-15 | 1-5 | Fluent | Fluent | Fluent | 2 | RESTART-DCM | No | No |
| RESTART_18 | T | Male | 11-15 | 1-5 | Fluent | Fluent | Fluent | 8 | RESTART-DCM | 4<sup>th</sup> degree relative | No |
| RESTART_19 | P | Male | 6-10 | 1-5 | Fluent | Fluent | Fluent | 20 | RESTART-DCM | No | No |
| RESTART_20 | T | Male | 11-15 | 1-5 | Fluent | Fluent | Fluent | - | Lidcombe program | No | No |
| RESTART_21 | P | Male | 6-10 | 1-5 | Mild | Mild to moderate | Mild to moderate | 27 | RESTART-DCM | No | No |
| RESTART_22 | T | Male | 11-15 | 1-5 | Fluent | Fluent | Fluent | 3 | Lidcombe program | No | Low intelligence, social/emotional problems, autism, ADD/ADHD |
| RESTART_23 | P | Male | 6-10 | 1-5 | Fluent | Fluent | Borderline | 19 | Lidcombe program | 4<sup>th</sup> degree relative | No |
| RESTART_24 | P | Male | 11-15 | 6-10 | Mild to moderate / moderate | Mild to moderate | Moderate to severe | 32 | RESTART-DCM | 4<sup>th</sup> degree relative (2x) | No |
| RESTART_25 | P | Male | 11-15 | 1-5 | Mild to moderate | Mild / mild to moderate | Mild / mild to moderate | 26 | Lidcombe program | No | No |
| RESTART_26 | T | Male | 11-15 | 1-5 | Borderline | Fluent | Borderline | 2 | RESTART-DCM | 3<sup>rd</sup> degree relative | No |
| RESTART_27 | T | Female | 6-10 | 1-5 | Borderline | Fluent | - | 2 | RESTART-DCM | 2<sup>nd</sup> degree relative | No |
| RESTART_28 | T | Male | 6-10 | 1-5 | Fluent | Fluent | Fluent | 4 | Lidcombe program | No | No |
| RESTART_29 | T | Female | 11-15 | 1-5 | Borderline | Fluent | Borderline | 2 | RESTART-DCM | 3<sup>rd</sup> degree relative | No |
| RESTART_30 | T | Female | 6-10 | 1-5 | Fluent | Fluent | Fluent | 2 | Lidcombe program | No | No |
| RESTART_31 | T | Female | 6-10 | 1-5 | Fluent | Fluent | Fluent | 2 | Lidcombe program | No | Hearing problems |
| RESTART_32 | P | Male | 11-15 | 1-5 | Mild to moderate | Mild | Mild | - | Lidcombe program | 4<sup>th</sup> degree relative | Dyslexia |
| RESTART_33 | P | Male | 11-15 | 1-5 | Moderate | Moderate | Moderate to severe | 33 | RESTART-DCM | No | No |
| RESTART_34 | A | Male | 6-10 | 1-5 | Fluent | Fluent | Fluent | 8 | Lidcombe program | No | Dyslexia |
| RESTART_35 | T | Female | 11-15 | 6-10 | Fluent | Fluent | Borderline case | 2 | RESTART-DCM | No | Problems with speech and language |
| RESTART_36 | P | Male | 11-15 | 6-10 | Fluent | Fluent | Fluent /<br>borderline | 15 | RESTART-DCM | No | Add/adhd |
| RESTART_37 | T | Female | 6-10 | 1-5 | Fluent | Fluent | Fluent | 0 | Lidcombe program | No | No |
| RESTART_38 | T | Male | 6-10 | 1-5 | Fluent | Fluent | Fluent | - | RESTART-DCM | No | No |
| RESTART_39 | T | Male | 6-10 | 1-5 | Fluent | Fluent | Fluent | 2 | Lidcombe program | No | No |
| RESTART_40 | T | Female | 6-10 | 1-5 | Fluent | Fluent | Fluent | 0 | RESTART-DCM | 2<sup>nd</sup> and 4<sup>th</sup> degree<br>relative | No |
| RESTART_41 | T | Male | 11-15 | 1-5 | Fluent | Fluent | - | - | Lidcombe program | 4<sup>th</sup> degree<br>relative | Dyslexia, ADD/ADHD |
| RESTART_42 | P | Male | 11-15 | 6-10 | Moderate | Mild to<br>moderate | Moderate | 28 | RESTART-DCM | 2<sup>nd</sup> and 3<sup>rd</sup> degree<br>relative | ADD/ADHD, Gilles de la<br>Tourette |
| RESTART_43 | T | Male | 6-10 | 1-5 | Fluent | Fluent | Borderline | - | Lidcombe program | 4<sup>th</sup> degree<br>relative | No |
| RESTART_44 | T | Male | 6-10 | 1-5 | Borderline | Borderline | Borderline | 2 | Lidcombe program | 2<sup>nd</sup> and 3<sup>rd</sup> degree<br>relative | No |
| RESTART_45 | A | Male | 10 | 1-5 | Borderline | Mild | Fluent | - | RESTART-DCM | No | Dyslexia |
| RESTART_46 | P | Male | 11-15 | 1-5 | Moderate | Moderate<br>to severe | Moderate<br>to severe | 32 | Lidcombe program | 3<sup>rd</sup> degree<br>relative | No |
| RESTART_47 | P | Male | 11-15 | 6-10 | Moderate | Mild | Borderline | 7 | Lidcombe program | No | No |
| RESTART_48 | A | Male | 6-10 | 1-5 | Fluent | Fluent | Fluent | 2 | RESTART-DCM | No | Dyslexia, ADD/ADHD |
| RESTART_49 | T | Female | 6-10 | 1-5 | Fluent | Borderline | - | 2 | Lidcombe program | 2<sup>nd</sup> degree<br>relative | Hearing problems |
| RESTART_50 | T | Male | 6-10 | 1-5 | Fluent | Fluent | Borderline | - | RESTART-DCM | 3<sup>rd</sup> degree<br>relative | Dyslexia |
| RESTART_51 | T | Male | 6-10 | 1-5 | Fluent | Fluent | Fluent | 3 | RESTART-DCM | No | Dyslexia |
| RESTART_52 | T | Female | 6-10 | 1-5 | Fluent | Fluent | Fluent | 2 | RESTART-DCM | 2<sup>nd</sup> degree<br>relative | Add/adhd |
| RESTART_53 | P | Male | 6-10 | 1-5 | Mild to<br>moderate | Moderate | Moderate | 28 | Lidcombe program | 2<sup>nd</sup> degree<br>relative | Dyslexia, lateralization<br>problems |
| RESTART_54 | P | Male | 6-10 | 1-5 | Mild | Mild to<br>moderate | Mild | 24 | RESTART-DCM | 4<sup>th</sup> degree<br>relative | Hearing problems |
| RESTART_55 | A | Male | 11-15 | 6-10 | Mild to<br>moderate | Moderate | Mild | 24 | RESTART-DCM | 4<sup>th</sup> degree<br>relative | No |
| RESTART_56 | P | Male | 6-10 | 1-5 | Borderline | Fluent | Fluent /<br>borderline | 23 | Lidcombe program,<br>speech/stutter therapy<br>(unknown method) | No | No |
| RESTART_57 | A | Male | 6-10 | 1-5 | Severe | Fluent | Mild | - | RESTART-DCM | No | Dyslexia |
| **MEGS_1** | P | Female | 6-10 | 6-10 | - | 5 | - | - | Speech/stutter therapy: (unknown method) + to learn speech techniques + to change negative thoughts and feelings | No | No |
| **MEGS_2** | P | Male | 6-10 | 1-5 | - | 7 | - | - | Speech/stutter therapy to learn speech techniques | Na | No |
| **MEGS_3** | P | Male | 6-10 | 1-5 | - | 8 | - | - | Speech/stutter therapy (unknown method); | 2<sup>nd</sup> degree relative | No |
| **MEGS_4** | P | Male | 6-10 | 1-5 | - | 2 | - | - | Speech/stutter therapy: (unknown method) + to learn speech techniques | No | No |
| **MEGS_5** | P | Male | 11-15 | 1-5 | - | 5 | - | - | Lidcombe program | 2<sup>nd</sup> degree relative | No |
| **MEGS_6** | P | Male | 6-10 | 1-5 | - | 5 | - | - | Speech/stutter therapy: (unknown method) + to learn speech techniques; Cognitive therapy | 2<sup>nd</sup> degree relative | No |
| **MEGS_7** | P | Female | 6-10 | 1-5 | - | 5 | - | - | Speech/stutter therapy: (unknown method) + to learn speech techniques | 2<sup>nd</sup> degree relative | No |
| **MEGS_8** | P | Male | 6-10 | 1-5 | - | 7 | - | - | Speech/stutter therapy: (unknown method) + to learn speech techniques; | 1<sup>st</sup> degree relative | No |
| **MEGS_9** | P | Male | 6-10 | 1-5 | - | 4 | - | - | Speech/stutter therapy: to learn speech techniques + to change negative thoughts and feelings + to say what you want to say without avoiding sounds/words/situations | No | No |
| **MEGS_10** | P | Male | 6-10 | 1-5 | - | 6 | - | - | Speech/stutter therapy (unknown method) | No | No |
| **MEGS_11** | P | Male | 11-15 | 1-5 | 7 | 7 | - | - | Hausdorfer method | No | No |
| **MEGS_12** | P | Female | 11-15 | 1-5 | 4 | 4 | - | - | Speech/stutter therapy (unknown method) | 2<sup>nd</sup> degree relative | No |
| **MEGS_13** | P | Male | 11-15 | 1-5 | 6 | 7 | - | - | Lidcombe therapy; Speech/stutter therapy to learn speech techniques | No | No |
| **MEGS_14** | P | Male | 11-15 | 1-5 | 3 | 4 | - | - | Lidcombe therapy; RESTART-DCM | No | Spelling difficulty |
| MEGS_15 | P | Male | 11-15 | 1-5 | 7 | 7 | - | - | Speech/stutter therapy (unknown method) + to learn speech techniques | 2<sup>nd</sup> (2x) and 3<sup>rd</sup> degree relative | No |
| MEGS_16 | P | Male | 11-15 | 6-10 | 4 | 4 | - | - | Speech/stutter therapy (unknown method) | 2<sup>nd</sup> degree relative | No |
| KST_1 | P | Male | - | - | - | - | - | - | - | No | - |
| KST_2 | P | Male | 26-30 | 1-5 | 6 | - | - | - | - | No | No |
| KST_3 | P | Female | 26-30 | 1-5 | 6 | - | - | - | - | No | No |
| KST_4 | P | Female | 46-50 | 6-10 | 7 | - | - | - | - | No | No |
| KST_5 | P | Male | 26-30 | 1-5 | 3 | - | - | - | - | 1<sup>st</sup> degree relative | No |
| KST_6 | P | Female | 26-30 | 1-5 | 4 | - | - | - | - | No | Otitis media |
| KST_7 | P | Female | 36-40 | 6-10 | 1 | - | - | - | - | 1<sup>st</sup> degree relative | No |
| KST_8 | P | Male | 41-45 | 1-5 | 5 | - | - | - | - | No | No |
| KST_9 | P | Male | 26-30 | 1-5 | 7 | - | - | - | - | No | Hay fever |
| KST_10 | P | Male | - | - | - | - | - | - | - | No | - |
| KST_11 | P | Male | - | - | - | - | - | - | - | No | - |
| KST_12 | P | Male | 36-40 | 1-5 | 5 | - | - | - | - | No | No |

A dash (-) indicates no data available. St: stuttering, classified as P: Persistent and T: transient. @Age in years, in age ranges (1-5; 6-10;11-15; etc). #numbers are on scale of 1 to 10, where 1 indicates no stuttering, and 10 indicates very severe stuttering (he/she can hardly produce a word). \*For RESTART: age during RESTART trial.

**Supplemental Table 2:**
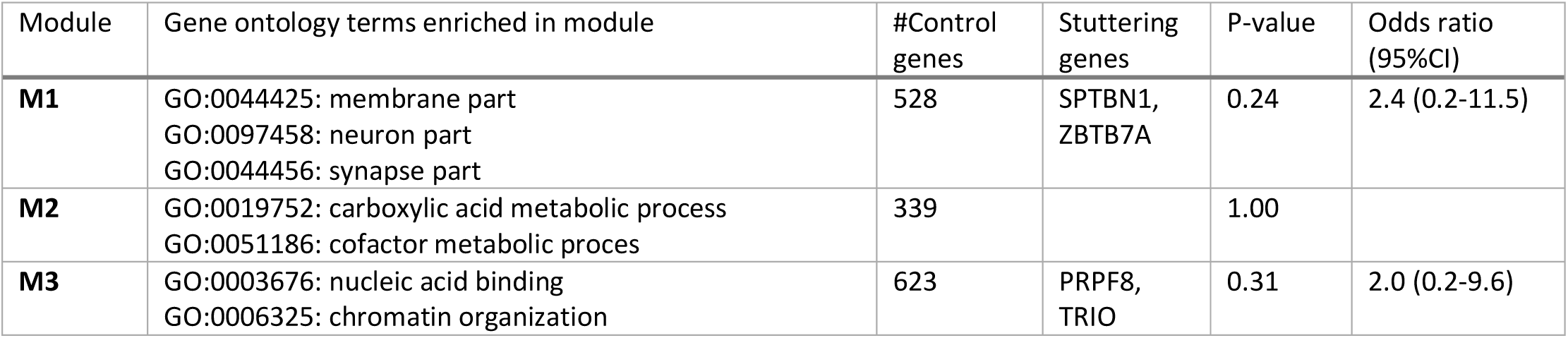
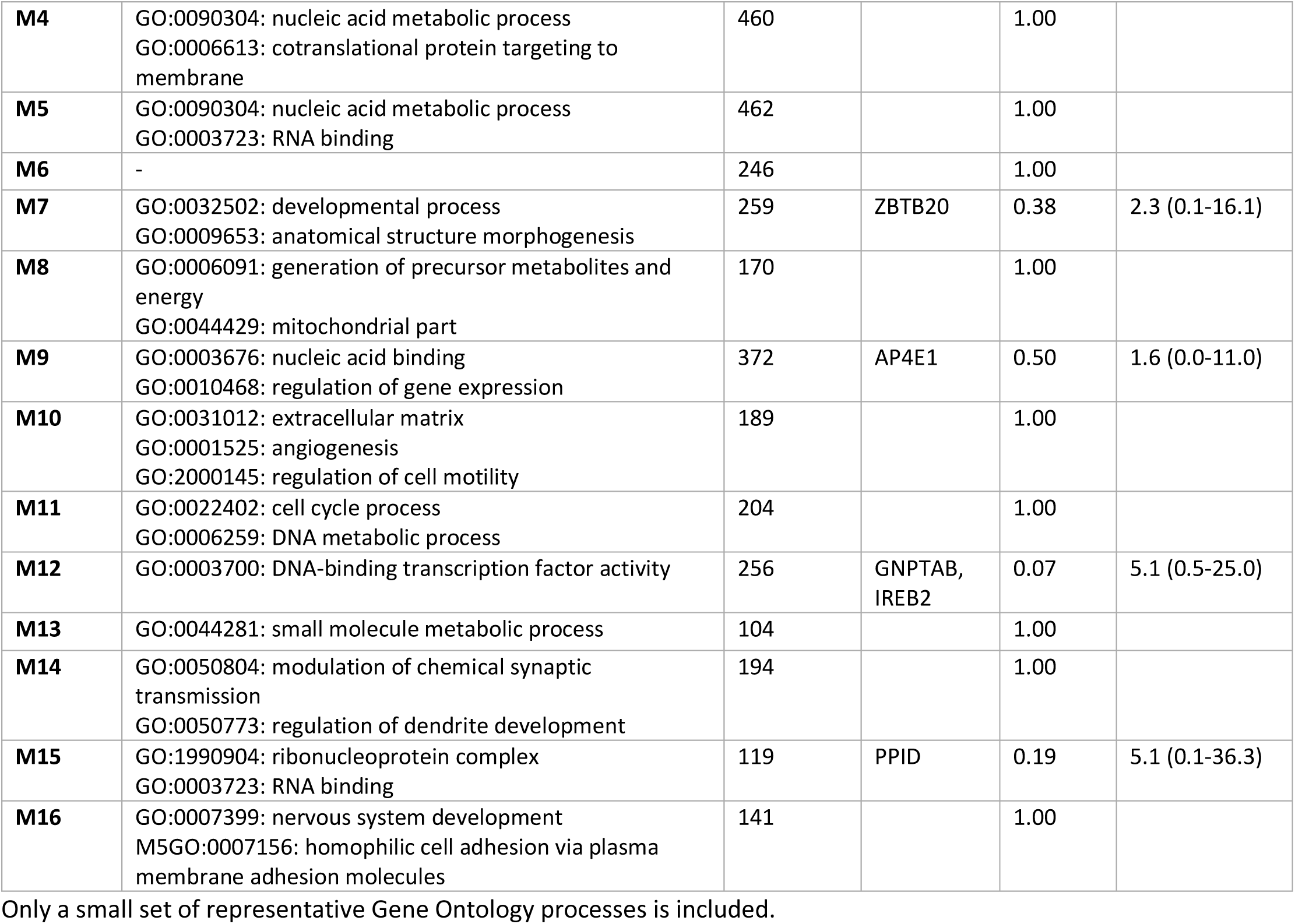
Enrichment analysis in developmental brain gene expression modules.

| Module | Gene ontology terms enriched in module | #Control genes | Stuttering genes | P-value | Odds ratio (95%CI) |
| --- | --- | --- | --- | --- | --- |
| **M1** | GO:0044425: membrane part<br>GO:0097458: neuron part<br>GO:0044456: synapse part | 528 | SPTBN1,<br>ZBTB7A | 0.24 | 2.4 (0.2-11.5) |
| **M2** | GO:0019752: carboxylic acid metabolic process<br>GO:0051186: cofactor metabolic proces | 339 |  | 1.00 |  |
| **M3** | GO:0003676: nucleic acid binding<br>GO:0006325: chromatin organization | 623 | PRPF8,<br>TRIO | 0.31 | 2.0 (0.2-9.6) |
| **M4** | GO:0090304: nucleic acid metabolic process<br>GO:0006613: cotranslational protein targeting to membrane | 460 |  | 1.00 |  |
| **M5** | GO:0090304: nucleic acid metabolic process<br>GO:0003723: RNA binding | 462 |  | 1.00 |  |
| **M6** | - | 246 |  | 1.00 |  |
| **M7** | GO:0032502: developmental process<br>GO:0009653: anatomical structure morphogenesis | 259 | ZBTB20 | 0.38 | 2.3 (0.1-16.1) |
| **M8** | GO:0006091: generation of precursor metabolites and energy<br>GO:0044429: mitochondrial part | 170 |  | 1.00 |  |
| **M9** | GO:0003676: nucleic acid binding<br>GO:0010468: regulation of gene expression | 372 | AP4E1 | 0.50 | 1.6 (0.0-11.0) |
| **M10** | GO:0031012: extracellular matrix<br>GO:0001525: angiogenesis<br>GO:2000145: regulation of cell motility | 189 |  | 1.00 |  |
| **M11** | GO:0022402: cell cycle process<br>GO:0006259: DNA metabolic process | 204 |  | 1.00 |  |
| **M12** | GO:0003700: DNA-binding transcription factor activity | 256 | GNPTAB,<br>IREB2 | 0.07 | 5.1 (0.5-25.0) |
| **M13** | GO:0044281: small molecule metabolic process | 104 |  | 1.00 |  |
| **M14** | GO:0050804: modulation of chemical synaptic transmission<br>GO:0050773: regulation of dendrite development | 194 |  | 1.00 |  |
| **M15** | GO:1990904: ribonucleoprotein complex<br>GO:0003723: RNA binding | 119 | PPID | 0.19 | 5.1 (0.1-36.3) |
| **M16** | GO:0007399: nervous system development<br>M5GO:0007156: homophilic cell adhesion via plasma membrane adhesion molecules | 141 |  | 1.00 |  |

Only a small set of representative Gene Ontology processes is included.

